# Cultural adaptation and content validity of a Chinese translation of the “Person-Centered Primary Care Measure”: findings from cognitive debriefing

**DOI:** 10.1101/2020.07.15.20154179

**Authors:** Tsui Yee Emily Tse, Lo Kuen Cindy Lam, King Ho Carlos Wong, Chin Weng Yee, Rebecca S. Etz, Stephen J. Zyzanski, Kurt C. Stange

**Affiliations:** Department of Family Medicine and Primary Care, Li Ka Shing Faculty of Medicine, The University of Hong Kong, 3/F, Ap Lei Chau Clinic, 161 Main Street, Ap Lei Chau, Hong Kong, China; Department of Family Medicine and Population Health, School of Medicine, Virginia Commonwealth University, 830 East Main Street, 6th floor, Room 629, Richmond, Virginia 23298-0101, The United States of America; Department of Family Medicine and Community Health, School of Medicine, Case Western Reserve University, Bolwell 1200, 11100 Euclid Ave, Cleveland, Ohio 44106, The United States of America

**Keywords:** Patient-Centered Care, Primary Health Care, Cognitive Debriefing, Validation Study

## Abstract

**Objectives:** To develop an equivalent Chinese translation of the Person-Centered Primary Care Measure (PCPCM) and to establish its cultural adaptability and content validity through cognitive debriefing.

**Design:** The original English PCPCM was first translated into Chinese by double forward-translation by professional translators. The reconciliated Chinese version was then doubly back-translated into English by two other professional translators blinded to the forward-translation. Upon affirmation on its linguistic equivalence with the developers of the original English PCPCM, the reconciliated Chinese PCPCM was sent for cognitive debriefing with twenty Chinese-speaking primary care subjects by a trained interviewer using structured probing questions to collect their opinions on the clarity, comprehensibility and relevance of each item and response option in the Measure.

**Setting:** Subjects were invited from a primary care clinic in Hong Kong to undergo the cognitive debriefing interviews. The interviews were divided into four groups chronologically to allow revision of the items to be made in between.

**Participants:** Ten males of age ranged from twenty-eight to sixty-eight and ten females of age ranged from thirty-seven to seventy completed the cognitive interviews. They were all Cantonese-speaking Chinese recruited by convenience sampling. Subjects with cognitive impairment, could not read Chinese, too old or too sick to complete the interviews were excluded from the study.

**Results:** An average of 3.3 minutes (range 3 to 4 minutes) was required for the subjects to self-complete the Measure. All items were generally perceived to be easily understood and relevant. Modifications were made to items with the content validity index on clarity or understanding <0.8 in each round of the interviews or if a majority of the subjects suggested rewording. Revisions were made to two items in the Chinese PCPCM throughout the whole cognitive debriefing process before the final version was confirmed. The average content validity index (CVI) on clarity of the Chinese PCPCM items ranged from 0.75 to 1. The average CVI on understanding ranged from 0.7 to 1. The average CVI on relevance ranged from 0.55 to 1.

**Conclusions:** The content validity of the PCPCM was good enough to allow further testing of its psychometric properties in a larger population.

**Key Points:** *Question:* Is our Chinese translation of The Person-Centered Primary Care Measure (PCPCM) a culturally adaptable and valid measure?

*Finding:* Our Chinese version of the PCPCM was confirmed to be culturally adaptable. It showed high content validity indices regarding its clarity, understanding and relevance through cognitive debriefing.

*Meaning:* This Chinese version of the PCPCM is ready for further testing of its psychometric properties in a larger population.

## Introduction

The Person-Centered Primary Care Measure (PCPCM) (Appendix 1) was developed in 2019 in the US with the aim to measure concisely the value of a primary care practice grounded in the experience of patients, clinicians and health care payers.(1) It is scored on a 4-point scale: definitely, mostly, somewhat and not at all. Patients need to be engaged in some information processing before responding to the questionnaire questions. They have to interpret the questions and retrieve their consultation experience as they fill out the questionnaire. They have to decide on their way of response and choose a response option which best fits them.(2-4) Subjects have to interpret the meaning of words or phrases in the questionnaire. Previous experience in the field revealed that translation itself (of questionnaires from a foreign language) may be a source of confusion for the respondents.(4, 5) When response options do not correspond to the subjects’ situations, they may become confused and do not know which response option to choose.(3, 4) Researchers need to look for the problems and correct them before the questionnaire can be formally administered in the general population.

In order for the PCPCM to be applicable to another culture, it has to be translated to the native language and confirmed to be valid in the target population. Ensuring the content validity in that target subjects’ interpretation of the questionnaire items being equivalent to what the original questionnaire developer intends to measure is a pre-requisite for further psychometric testing. Moreover, the response options of each item need to allow the subjects to respond in the way which best fits their opinions and situations.

The National Center for Health Statistics Questionnaire Design Research Laboratory at the Centers for Disease Control and Prevention advises adopting cognitive debriefing to identify any problem or confusion in questionnaires.(3) In cognitive debriefing, interviewers apply one-on-one interviews to investigate the approach subjects employed to process the data when they answer the questions. Problems in item interpretation, decision processes, and response option selection can be recognized. Other problems, for instance, instructions, design and structure of the questionnaire can also be identified through cognitive debriefing.(3, 4)

This paper describes our first step to adapt the PCPCM for the evaluation of patient-centered care in primary care in Hong Kong where 95% of the population are Chinese. The aim of this study was to establish the cultural adaptability and content validity of a Chinese version of the PCPCM. The objectives were to develop an equivalent Chinese translation of the PCPCM, and to evaluate the clarity, understanding and relevance of each item. This will in turn provide an equivalent Chinese PCPCM that is applicable to Chinese primary care patients for pilot psychometric testing.

## Methodology

### Development of the Chinese PCPCM and evaluation of content validity

The Chinese translation of the PCPCM was developed according to the International Society For Pharmacoeconomics and Outcome Research (ISPOR) Principles of Good Practice: The Cross-Cultural Adaptation Process for Patient-Reported Outcomes Measures (6). At the ‘Preparation’ stage, an Expert Review Panel consisting of six local primary care experts were invited to assess the face validity of the original English PCPCM in the Hong Kong Chinese context. They unanimously agreed that the PCPCM was measuring the important aspects of primary care including ‘accessibility’, ‘comprehensiveness’, ‘community-based’, ‘continuity of care’, ‘holistic care’, ‘coordinated care’, ‘evidence-based practice’, ‘rapport building’, ‘patient advocate’, ‘preventive care’, ‘patient enablement’ and ‘patient-centered care’. They confirmed no amendment was needed for the PCPCM prior to translation.

Two professional translators who are native Chinese speakers, were employed to translate the original English version of the PCPCM into Chinese independently. Two bilingual investigators (ETYT and CLKL) reviewed the translations and formed the first draft of the Chinese PCPCM. Another two professional translators blinded to the original PCPCM were employed to back-translate the first draft to English. The back-translation was assessed and confirmed to be equivalent to the original measure by its developers (RE and KS). This first draft of the Chinese translation (Appendix 2) was sent for cognitive debriefing with twenty Chinese patients attending a public sector primary care clinic in Hong Kong to evaluate the clarity and interpretation of each item and response option.

### Sampling of subjects

Subjects were recruited from a government-funded primary care clinic in Hong Kong where nearly all subjects were Cantonese-speaking Chinese. Subject inclusion criteria were Cantonese-speaking adults (≥ 18 years old) without cognitive impairment and able to read Chinese. Exclusion criteria were subjects who were too old or too sick to complete the interview. The sampling was purposive to include subjects with a wide range of ages and education levels with an equal distribution of gender.

### Procedures

The cognitive debriefing was conducted between July to August 2019. A trained research assistant carried out the cognitive debriefing using an interviewer guide with structured probing questions (**Table 1**). Subjects were encouraged to give comments on any difficulty in completing the questionnaire and give recommendations to replace any unclear wording. All the debriefing interviews were conducted one-on-one in the primary care clinic. Written informed consent was obtained prior to each interview.

**Table 1.** Structured Cognitive Debriefing Interviewer Guide.

| Purpose | Probing Question |
| --- | --- |
| Determine roughly the subjects' comprehension of the questionnaire and obtain comments on the items, response options and the questionnaire design in general. | G1. Are the questions generally clear, easy to understand, easy to answer? |
|  | G2. Overall, is the measure relevant to your situation? (Are the items meaningful and important to you?) |
|  | G3. Are the instructions clear and easy to understand? |
|  | G4. Is the format easy to follow? (Is it easy to complete on your own?) |
| Determine if the items or response options are confusing or problematic. | 1. Did you have any difficulty understanding this item /response |
|  | scale? |
| Determine if subjects perceive the items the same or similar as the developer's intention. | 2. What does this item mean to you? |
| Identify any confusing words or phrases. | 3. Would you reword this item? (If so, how would you reword it?) |
| Determine if the items are relevant to the subjects. | 4. Is this item relevant to your situation? |
| Determine if the subjects can easily choose a response option that best fits their situation. | 5. Are the response options consistent with this item? (If no, please explain the difficulty and suggest how you would reword them.) |

At the start, the interviewer explained the aim of the study and the procedures of the cognitive debriefing to the subject. The interview was audiotaped. Demographic data (**Table 2**) of the subject were collected. The subject then completed the Chinese PCPCM by him/herself and the time of completion was recorded. The audiotaping and cognitive debriefing started afterwards. The subject answered four questions on their general impression on the questionnaire, and then five probing questions for each item of the PCPCM. Each cognitive debriefing interview lasted 20 to 30 minutes. Each subject was given HKD100 (∼USD13) supermarket voucher in appreciation of his/her contribution to the study.

**Table 2.** Characteristics of cognitive debriefing subjects.

| Demographic Information | N=20, (%) |
| --- | --- |
| **Gender** |  |
| Female | 10 (50%) |
| Male | 10 (50%) |
| **Age (years)** |  |
| Mean | 55.35 |
| Range | 28-70 |
| **Education** |  |
| Not educated / before primary school | 1 (5%) |
| Primary school | 2 (10%) |
| Junior Secondary School | 3 (15%) |
| Higher Secondary School | 8 (40%) |
| Post-secondary colleges<br>(non-degree program) | 5 (25%) |
| Universities / master or above | 1 (5%) |
| **Employment** |  |
| Employed | 10 (50%) |
| Housewife | 2 (10%) |
| Unemployed / retired | 8 (40%) |

The subjects’ answers to the interview were recorded and transcribed verbatim. Interview results were summarized in a tabular format. The transcript was reviewed by the investigators to identify any problem in the content of the draft Chinese PCPCM after the completion of each round of interviews. When a problem was recognized, the investigators (ETYT and CLKL) deliberated on the problem item and revised the content accordingly, and further tested in the next group of subjects. The process continued until there was no more problem found in each item. The whole cognitive debriefing process was performed through 4 rounds of interviews with 20 different subjects. The first, second, third and fourth rounds consisted of 9, 5, 3 and 3 subjects respectively. The content validity index (CVI) was calculated by the total number of positive ratings divided by the number of subjects in that round. Revisions were made to items with the content validity index on clarity or understanding <0.8 after each round of interview or if a majority of the subjects suggested a rewording. The revised measure was subsequently tested with the next round of subjects until no more problem was identified (at the fourth round).

## Results

The mean completion time of the Chinese PCPCM amongst the 20 subjects was 3.3 minutes (ranged from 3 to 4 minutes).

The eleven items related to person-centeredness together with the response scale, and an additional item asking for the duration of the subject having known the doctor (Appendix 2) underwent content validation. As mentioned in the methodology section, the whole cognitive debriefing process was performed through 4 rounds of interviews with 20 different subjects. Sixteen subjects (80%) commented the items in the Chinese PCPCM in general were clear, easy to understand and to answer (question G1 stated on **Table 1**). All subjects confirmed relevance (question G2 stated on **Table 1**) and clarity of the instructions (question G3 stated on **Table 1**) of the Measure on the whole. One out of the 20 subjects commented on the format of the response scale (question G4 stated on **Table 1**): He found the distinction between ‘mostly’ and ‘somewhat’ to be unclear. He suggested that could be changed to a percentage scale to indicate the respondent’s degree of agreement with the item. Another subject (an elderly aged 68) expressed that it was a bit difficult to complete the Chinese PCPCM on his own.

After obtaining the subjects’ general impression on the Chinese PCPCM, they were asked to explain the meaning of each item and to suggest if any rewording needed to improve comprehension. The average CVIs on clarity, understanding and relevance of each item are shown in **Table 3**. As revealed by the answers to the general probing questions, majority of patients actually found most of the question items to be clear, easy to understand, relevant to them and did not require rewording. The exception was for items five, eight, nine and ten.

**Table 3.** Average Content Validity Index (CVI) on clarity, understanding and relevance of each item in the PCPCM during the 4 rounds of cognitive debriefing interviews.

| Item | CVI |  |  |
| --- | --- | --- | --- |
|  | Clarity | Understanding | Relevance |
| 1. The practice makes it easy for me to get care. | 1 | 1 | 0.95 |
| 2. This practice is able to provide most of my care. | 1 | 1 | 0.95 |
| 3. In caring for me, my doctor considers all factors that affect my health. | 0.95 | 0.95 | 1 |
| 4. My practice coordinates the care I get from multiple places. | 0.95 | 0.95 | 0.95 |
| 5. This doctor or practice knows my needs in all aspects. | 0.85 | 0.90 | 0.95 |
| 6. My doctor and I have been through a lot | 0.95 | 0.95 | 0.95 |
| together. |  |  |  |
| 7. My doctor or practice stands up for me. | 0.95 | 0.95 | 1 |
| 8. The care I get takes into account knowledge of my family. | 1 | 1 | 0.55 |
| 9. The care I get in this practice is informed by knowledge of my community. | 0.95 | 1 | 0.55 |
| 10. Over time, this practice helps me to meet my health-related goals. | 0.75 | 0.70 | 0.85 |
| 11. Over time, my practice helps me stay healthy. | 1 | 1 | 0.90 |
| 12. How many years have you known this doctor | 1 | 1 | 1 |
| 13. Response scale: Definitely/ Mostly/ Somewhat/ Not at all | 1 | 0.90 | 1 |

For item five, the English version was ‘This doctor or practice knows me as a person.’ We translated that into ‘這位醫生或這間診所對我個人很了解’ initially. This translation literally means ‘This doctor or practice understands me well.’ Although the CVI on clarity and understanding in the first round of interviews with the nine subjects was 0.89 and 1 respectively (**Table 4**), subjects actually interpreted the meaning quite diversely. For example, they suggested the meaning to be knowing his or her medical background, habits, diet pattern, drug allergies, etc. In view of the broad interpretation spectrum, some subjects suggested a rewording to limit the scope to ‘medical aspect’. Upon consulting the original PCPCM developers in the US, they confirmed that it was their intention to allow the subjects to have their own interpretations because how primary care had functioned and had added value to patients’ lives were actually complex notions. They suggested us to reword the translation to cover a larger meaning including the patient’s day to day life, medical problems, risk factors, health behaviors, what is important in his or her life, the patient’s dreams, failures and even larger aspirations. We hence reworded the question to ‘這位醫生或這間診所對我全人很了解’ meaning ‘This doctor or practice knows me as a person holistically’ in the second round of interviews. In contrary to our expectation, the CVI on clarity and understanding did not increase but dropped to 0.6. The subjects commented that the word holistic was remote and inaccurate in their relationship with the doctors. From their experience, doctors would not know too much about a patient’s life other than the medical aspect. The local investigators (ETYT and CLKL) deliberated on the item and suggested a rewording to ‘這位醫生或這間診所對我各方面的需要都很了解’ meaning ‘This doctor or practice knows my needs in all aspects’ as this seemed to be more comprehensible within the local Chinese context. The suggestion was supported by the US team. Upon testing that out with the subjects in the third and fourth rounds of interviews, the CVI on clarity and understanding rose to 1.

**Table 4.**
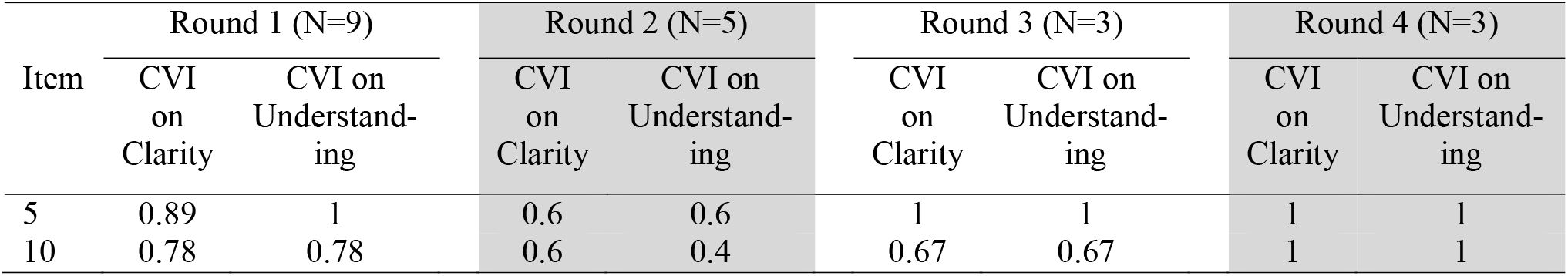
CVI on clarity and understanding of items 5 and 10 in each round of interviews.

The average CVI on relevance of items eight and nine were both 0.55. Those items were ‘The care I get takes into account knowledge of my family’ and ‘The care I get in this practice is informed by knowledge of my community’. For item eight, a few of the subjects stated that they had never talked about their family in front of the doctors. They thought that was beyond the scope of medical consultations. One subject even expressed that being asked about the family background during medical consultations would be too intrusive into one’s privacy. Amongst many subjects who thought the item was irrelevant to them, they believed it would be of higher relevance to patients at advanced age or with physical disabilities. For item nine, some subjects pointed out that the doctors in this clinic might not actually know much about the local community as they were not living in this district. Even if they knew the district well, that had nothing to do with taking care of patients. Only a minority of the subjects made the link that the doctor or the practice could mobilize resources in the community to help patients or could direct patients to services in the vicinity.

For item ten, the average CVI on clarity and understanding was 0.75 and 0.70 (**Table 3**) respectively. The English version of this item was ‘Over time, this practice helps me to meet my goals.’ We translated that into ‘這間診所一直在幫助我實現我的目標’ initially. The CVI on clarity and understanding in the first round of interviews with the nine subjects was both 0.78 (**Table 4**). When looking into the subjects’ comments in details, actually many of them interpreted the item within the context of ‘health-related goals’. As the intention of this item in the original English PCPCM was to explore a larger context of different goals in life and the CVIs were just marginally low, we tried to keep the translation to test through the second and third round of interviews. However, the CVI on clarity still remained low (0.6 and 0.67) in these two rounds whilst the CVI on understanding further dropped to 0.4 and 0.67 in the second and third round respectively. Advice was sought from our US team again and they agreed that it would be appropriate to add ‘health-related’ to the ‘goals’ concerned. We hence reworded the question to ‘這間診所一直在幫助我實現我健康相關的目標’ meaning ‘Over time, this practice helps me to meet my health-related goals’ in the fourth round of interviews. This time, the CVI on clarity and understanding rose to 1 (**Table 4**).

Interpretations on individual items, suggestions of rewording by subjects and the follow up actions taken by the investigators (where applicable) are listed in **Table 5**.

**Table 5.** Subjects’ interpretations on individual item, suggested rewording and investigators’ follow up actions.

| Item | Subjects’ interpretation of items, suggested rewording and follow up actions |
| --- | --- |
| 1 | Subjects understood the item and most of them correlated it with geographical accessibility, the phone booking system and the professional services provided. |
| 2 | All subjects understood the item and correlated it with the context in “general practice”. |
| 3 | All subjects understood the item. They correlated the ‘factors affecting their health’ to a broad range of contexts including their symptoms, medical records, the investigations needed, complications of diseases, well-being on |
|  | the whole, psychological factors, habits, drug usage and diet pattern, etc. |
| 4 | Most subjects showed understanding to it. They commonly linked the item to making referrals to other specialties or allied health services. Some correlated it with services suggested elsewhere (for example, wound dressing initiated by the emergency department). |
| 5 | In the first round of interviews (N = 9), subjects interpreted the meaning of ‘understands me well’ quite diversely. Their interpretations included understanding his or her medical background, habits, diet pattern, drug allergies, etc. After reworded to ‘knows me as a person holistically’ in the second round (N = 5), the subjects thought that “holistic” was too general to be real in their experience. Further deliberation of the item was made amongst the local and US investigators. The item was subsequently rephrased as “knows my needs in all aspects”. No more question was raised in the clarity and understanding of the item in the third (N = 3) and fourth (N = 3) rounds of interviews. However, some subjects suggested changing the words “This doctor or practice” to “The doctors of this practice” as they might not be seeing the same doctor every time. The investigators decided there was no need to further change the translation. |
| 6 | Most of the subjects thought it was not their experience with the doctors in this clinic. The main reason was that they might not be seeing the same doctor every time. One subject appreciated this question as focusing on chronic diseases management by the same doctor. The investigators concluded that we should keep the translation unchanged. |
| 7 | Most subjects suggested that it should be made more specific in “stands up for me” in which aspect. They suggested fields like “confidentiality”, “the right to receive medical care”, and “putting patients’ benefits first”, etc. However, the investigators concluded we should keep the original translation to avoid narrowing down too much and running the risk of losing those important functions of primary care services in the subjects’ notions. |
| 8 | All subjects showed understanding to this item. However, many of them found it was not applicable to their situations. A subject suggested adding a response option of “Not Applicable” to the answers. The investigators concluded that we should keep the translation and response options unchanged. |
| 9 | Two subjects commented that the item was slightly unclear. Some of the subjects interpreted it as “knowledge of the community resources available” whilst some others interpreted it as “the general health or socio-economic condition of the community”. A few of them thought that the knowledge of the community was irrelevant to them. A subject suggested adding a response option of “Not Applicable” to the answers. The investigators decided to keep the translation and response options unchanged because the problem actually |
|  | stemmed from lack of experience by the subjects to the item. |
| 10 | Majority of the subjects commented that the word “goals” was not specific. They usually interpreted that as “health-related” goals and suggested adding these words to make the meaning more explicit. In the original English PCPCM, the “goals” actually refer to goals in a larger context. After thorough discussion amongst the investigators, we agreed that it was justifiable to add “health-related” to make the “goals” more comprehensible in the Chinese patients’ context. |
| 11 | All subjects commented this item was clear. |
| How many years have you known this doctor? | All subjects commented this item was clear. |
| Response options | 18 out of 20 subjects commented the response options were clear and selectable. Two subjects found the response options being unclear in the distinction between ‘mostly’ and ‘somewhat’. Both of them suggested that the response could be changed to a percentage scale to indicate the degree of agreement with the item instead of using categorical options.<br>A few subjects made further suggestions to the response options despite agreeing that the current choices were acceptable:<br>-One subject suggested that the response options should be reworded as “very satisfied”, “satisfied”, “dissatisfied” and “very dissatisfied”.<br>-One subject commented there should be an additional ‘neutral’ option to make the negative and positive options more balanced.<br>-Two subjects commented that for item 8 and item 9, an option of “not applicable” could be added.<br>The investigators concluded that the response options need not be changed. |

### Overall revisions made to the draft Chinese PCPCM

Based on the results of the cognitive debriefing interviews and discussion amongst the local and US investigators, revisions were made to items 5 and 10 only. The final version of the Chinese PCPCM is attached as Appendix 3.

## Discussion

A measure that can capture the patient-perceived value of primary care is much needed to evaluate the quality of care and to document the health benefit of interventions. The PCPCM is a standardized and valid solution to assess primary care practice from an individual’s perspective and was grounded in the experience of patients, clinicians and health care payers.(1) Our study showed that the concept was applicable to the Chinese culture and an equivalent Chinese translation was possible. The average content validity (CVI) index on clarity of each item is over 0.8 except item ten (0.75). The average CVI on understanding of each item is over 0.8 except item ten (0.7). The CVI in clarity and understanding of this item eventually reached 1 in the final round of interviews. The CVI on relevance of each item was >=0.85 except items eight and nine. All items and response scale were considered generally applicable and valid in the Chinese primary care subjects.

We found our subjects had different interpretations of the meaning of item 5. The idea that “The doctor or a practice knows me as a person” was rather foreign to the subjects attending busy public primary care clinics in Hong Kong. Some subjects interpreted it as “(The doctor) knows my medical record or my health conditions”. This demonstrated that semantic equivalence is not sufficient in the translation of a psychometric measure from one language to another. It is important to take into consideration of the cultural and contextual differences between the original and target populations. The clinical practice in US is different from that in Hong Kong. The primary care home model advocated in the US in the past two decades promoted more time on caring the patients on the whole and more attention to all aspects of their living.(7) Primary care in Hong Kong, similar to those in most other Chinese and Asian societies, is mainly doctor-led and the high workload limited the amount of time and scope of service patients can get. Another common problem in the system is that patients may not see the same doctor each time they attend because there are more than 10 doctors working in rotation in one clinic (as a norm in the local public primary care system). This leads patients to think along the line of different doctors “read and know their medical records or their health conditions” instead of “know them as a person”. Our study revealed interesting cultural differences in the expectation of person-centeredness within the context of primary care.

## Strengths and Limitations

In our study, formal double forward- and backward-translations were applied. The original authors of the PCPCM reviewed the English back-translation to assure semantic equivalence of the Chinese translation.

However, our study shared the same limitation with other studies using cognitive debriefing: Subjects may not have given “sufficient mental effort” to the debriefing and it is difficult to assess if they have.(8, 9) Subjects might just want to give a socially desirable response, i.e. faking good(5, 8) leading to futile results. Another problem is the potential danger of using probing questions to identify subjects’ comprehension problems. For simple questions, subjects may be so automatic to give responses that do not need much cognitive processes. If subjects are prompted for elaboration of the questions or recommendations of re-wording when none is available in their head, they may compose a vague reply rather than replying they have no idea.(8)

## Future Research

A respondent debriefing can be included in the cognitive debriefing. Other than just asking the probing questions on each item in the questionnaire, additional probing questions can also be asked on why the subjects chose the particular response items. It helps us to further understand how subjects interpreted the questions and how they reached their answers.(10-12) This kind of debriefing can help to recognize questions which subjects could not answer precisely.(10, 13)

## Conclusion

In search of a concise and comprehensive new measure to evaluate the value of a primary care practice from the patients’ perspective, the Chinese translation of the Person-Centered Primary Care Measure (PCPCM) is now available and ready for further psychometric testing on a wider population to confirm its validity, reliability, sensitivity and responsiveness. Eventually we can include patient-centered care as a routine measure of quality and outcome of primary care.

## Data Availability

All data relevant to the study are available upon reasonable request.

## Author contributions

The authors contributed to the concept of the study, analysis and interpretation of the data, drafting and critical revision of the manuscript for important intellectual content. The authors approved the final version for publication and took responsibility for the accuracy and integrity of the study.

## Special Acknowledgement

We would like to thank Professor Samuel YS Wong, Dr Lee Siu Yin Ruby, Dr Tsang Chiu Yee Luke, Dr Wan Wing Fai, Dr Chiu Chi Fai Billy (from the academic, professional body, private and public clinical service fields) for their contributions in the Expert Review Panel at the beginning of the study to confirm the face validity of the English PCPCM in our local context. Thanks also go to Ms. Lam Sau Mei Joyce, our project’s research assistant, who made a significant contribution to the implementation of the study, collection of the research data, statistical analysis, results interpretation and manuscript drafting.

## Declaration

### Ethical approval

HKU/HA HKW IRB reference number UW 18-492. Conflict of interest: none.

## Appendix 1. English version of the Person-Centered Primary Care Measure

**‘The Person-Centered Primary Care Measure’**

**Measuring what Matters in Primary Care**

Please circle the response that best fits your experience for each item. Thank you.

| PATIENT’S GENERAL ASSESSMENT OF TODAY’S VISIT | RESPONSE |
| --- | --- |
| The practice makes it easy for me to get care. | Definitely Mostly Somewhat Not at all |
| This practice is able to provide most of my care. | Definitely Mostly Somewhat Not at all |
| In caring for me, my doctor considers all factors that affect my health. | Definitely Mostly Somewhat Not at all |
| My practice coordinates the care I get from multiple places. | Definitely Mostly Somewhat Not at all |
| This doctor or practice knows me as a person. | Definitely Mostly Somewhat Not at all |
| My doctor and I have been through a lot together. | Definitely Mostly Somewhat Not at all |
| My doctor or practice stands up for me. | Definitely Mostly Somewhat Not at all |
| The care I get takes into account knowledge of my family. | Definitely Mostly Somewhat Not at all |
| The care I get in this practice is informed by knowledge of my community. | Definitely Mostly Somewhat Not at all |
| Over time, this practice helps me to meet my goals. | Definitely Mostly Somewhat Not at all |
| Over time, my practice helps me stay healthy. | Definitely Mostly Somewhat Not at all |
| PLEASE TELL US A BIT ABOUT YOURSELF |  |
| How many years have you known this doctor? | _____ (number of years) |

## Appendix 2. Initial Chinese version of the Person-Centered Primary Care Measure to undergo cognitive debriefing

**以人為本的基層醫療服務量表**

**量度以人為本基層醫療服務的重要元素**

請圈出下列句子中最符合您經驗的答案。謝謝。

| 患者對今天求診的整體評價 | 答案 |
| --- | --- |
| 這間診所讓我能輕易地得到醫療服務。 | 絕對正確 大部分正確 有點正確 完全不正確 |
| 這間診所能夠提供大部分我所需要的醫療服務。 | 絕對正確 大部分正確 有點正確 完全不正確 |
| 在照顧我時，我的醫生會考慮所有影響我健康的因素。 | 絕對正確 大部分正確 有點正確 完全不正確 |
| 這間診所能協調我在多個地方所接受的醫療服務。 | 絕對正確 大部分正確 有點正確 完全不正確 |
| 這位醫生或這間診所對我個人很了解。 | 絕對正確 大部分正確 有點正確 完全不正確 |
| 我和我的醫生共同經歷了很多健康的大小問題。 | 絕對正確 大部分正確 有點正確 完全不正確 |
| 我的醫生或診所維護我的權益。 | 絕對正確 大部分正確 有點正確 完全不正確 |
| 這裏的醫護人員會考慮到我的家庭情況從而對我作出適切的照顧。 | 絕對正確 大部分正確 有點正確 完全不正確 |
| 這裏的醫護人員認識我所屬的社區，從而給我適切的照顧。 | 絕對正確 大部分正確 有點正確 完全不正確 |
| 這間診所一直在幫助我實現我的目標。 | 絕對正確 大部分正確 有點正確 完全不正確 |
| 這間診所一直在幫助我維持健康。 | 絕對正確 大部分正確 有點正確 完全不正確 |
| 請告訴我們一些您的資料 |  |
| 您認識這位醫生多少年了？ | _____（年） |

## Appendix 3. Final Chinese version of the Person-Centered Primary Care Measure

**以人為本的基層醫療服務量表**

**量度以人為本基層醫療服務的重要元素**

請圈出下列句子中最符合您經驗的答案。謝謝。

| 患者對今天求診的整體評價 | 答案 |
| --- | --- |
| 這間診所讓我能輕易地得到醫療服務。 | 絕對正確 大部分正確 有點正確 完全不正確 |
| 這間診所能夠提供大部分我所需要的醫療服務。 | 絕對正確 大部分正確 有點正確 完全不正確 |
| 在照顧我時，我的醫生會考慮所有影響我健康的因素。 | 絕對正確 大部分正確 有點正確 完全不正確 |
| 這間診所能協調我在多個地方所接受的醫療服務。 | 絕對正確 大部分正確 有點正確 完全不正確 |
| 這位醫生或這間診所對我各方面的需要都很了解。 | 絕對正確 大部分正確 有點正確 完全不正確 |
| 我和我的醫生共同經歷了很多健康的大小問題。 | 絕對正確 大部分正確 有點正確 完全不正確 |
| 我的醫生或診所維護我的權益。 | 絕對正確 大部分正確 有點正確 完全不正確 |
| 這裏的醫護人員會考慮到我的家庭情況從而對我作出適切的照顧。 | 絕對正確 大部分正確 有點正確 完全不正確 |
| 這裏的醫護人員認識我所屬的社區，從而給我適切的照顧。 | 絕對正確 大部分正確 有點正確 完全不正確 |
| 這間診所一直在幫助我實現我健康相關的目標。 | 絕對正確 大部分正確 有點正確 完全不正確 |
| 這間診所一直在幫助我維持健康。 | 絕對正確 大部分正確 有點正確 完全不正確 |
| 請告訴我們一些您的資料 |  |
| 您認識這位醫生多少年了？ | _____（年） |

